# Validation of Emotional Stimuli Flashcards for Conducting ‘Response to Reward’ fMRI study among Malaysian students

**DOI:** 10.1101/2020.01.17.20017202

**Authors:** Nisha Syed Nasser, Hamed Sharifat, Aida Abdul Rashid, Suzana Ab Hamid, Ezamin Abdul Rahim, Mazlyfarina Mohamad, Rohit Tyagi, Siti Irma Fadhilah Ismail, Ching Siew Mooi, Subapriya Suppiah

**Affiliations:** Centre for Diagnostic Nuclear Imaging, Faculty of Medicine and Health Sciences, Universiti Putra Malaysia, Malaysia; Department of Imaging, Faculty of Medicine and Health Sciences, Universiti Putra Malaysia, Malaysia; Department of Diagnostic Imaging and Radiotherapy Programme, Faculty of Health Sciences, Universiti Kebangsaan Malaysia, Malaysia; Aerobe Pte. Ltd, Singapore; Department of Psychiatry, Faculty of Medicine and Health Sciences, Universiti Putra Malaysia, 43400 Serdang, Selangor, Malaysia; Department of Family Medicine, Faculty of Medicine and Health Sciences, Universiti Putra Malaysia, 43400 Serdang, Selangor, Malaysia

**Keywords:** Addiction, Affective ratings, Cravings, Picture database, Reward

## Abstract

Problematic Instagram Use (PIGU) is a specific-Internet-addiction disorder observed among the youth of today. fMRI, is able to objectively assess regional brain activation in response to addiction-specific rewards, e.g. viewing picture flashcards. Pictures uploaded onto Instagram by PIGUs have often been associated with risky behaviours in their efforts to gain more ‘Likes’, thus it is hypothesized that PIGUs are more drawn to ‘Negative-Emotional’ cues. To date, there is no local database with addiction-specific cues.

**Objective:** To conduct an out-of-scanner validation study to create a database of pictures using ‘Negative-Emotional’ cues that evoke responses of arousal among PIGUs.

**Method:** Forty-four Malaysian undergraduate students (20 PIGUs, 24 controls) were randomly recruited based on the evaluation using the Smartphone-Addiction-Scale-Malay version (SAS-M) and modified Instagram Addiction Test (IGAT); and fulfilled Lin et al. (2016) definition of addiction. They were administered 200 content-specific pictures that were multidimensional i.e. arousal (excitation/relaxation effects), approach-avoidance (motivational direction) and emotional valence (positive/negative feelings) to describe the PIGUs perception of the psychological properties of the pictures using a 9-point Likert scale.

**Results:** PIGUs viewing ‘Negative-Emotional’ cues demonstrated significant positive correlations between arousal & valence (z = 4.834, p < .001, effect size = 0.69) and arousal & avoidance-approach (z = 4.625, p < .001, effect size= 0.66) as compared to controls and were more frequently aroused by ‘Negative-Emotional’ type of stimuli.

**Conclusion:** A database of validated, addiction-specific pictures can be developed to potentiate any future cue-induced response to reward fMRI studies to assess PIGU.

## Introduction

Problematic Instagram Use (PIGU) is a behavioural addiction (BA) and considered as a type of specific-Internet-addiction, which is associated with cravings and overdependence of using Instagramsocial networking application (SNA) despite experiencing deterioration in activities of daily living among the addicted individual (Billieux 2012; Brand et al. 2016). To be precise, addiction is defined as a compulsive behaviour associated with continued indulgence in pleasure-seeking activities with no desire to give up, despite the detrimental effects (Volkow et al. 2016). Addiction is classically recognized as substance use disorders (SUDs) such as addiction to cannabis, tobacco, alcohol, hallucinogens, sedatives, caffeine, opioids, hypnotics, and anxiolytics; but also encompasses abnormal behaviours such as pathological gambling, preoccupation with the Internet, online gaming, and social networking (American Psychiatric Association 2013).

Instagram use among Malaysians has increased exponentially in the past decade giving rise to problematic behaviours related to its use (Khalid et al. 2018). Most studies have relied on questionnaire-based assessment e.g. the Smartphone Addiction Scale-Malay version (SAS-M) and modified Instagram Addiction Test (IGAT) (Ching et al. 2015; Syed Nasser et al. 2019) to establish commonalities between PIGU and other BA and SUDs, albeit lacking in neurobiological evidence to support the hypothesis. Advantageously, functional magnetic resonance imaging (fMRI) neuroimaging technique can objectively assess regional brain activations in response to addiction-specific rewards. In addition to using the questionnaire-based scoring criteria, PIGU is defined by a set of modified diagnostic criteria for smartphone addiction, which is an objective assessment as proposed by Lin et al. (2016).

PIGU can be defined as a modification of the proposed diagnostic criteria for smartphone addiction by replacing the word “the Smartphone” with the word “Instagram” that gives the sensitivity, specificity and diagnostic accuracy of 79.4%, 87.5%, and 84.3% respectively. The criteria comprise of three parts occurring within a 3-month period, namely (1) Criterion A: presence of ≥ 3 out of six symptoms (Repeated inability to resist using the Smartphone, Withdrawal symptoms as evidenced by dysphoric mood, irritability and anxiety when away from the smartphone for any period of time, Prolonged usage of the Smartphone for durations longer than initially intended, Unsuccessful attempts to reduce the Smartphone usage, Excessive preoccupations with using the Smartphone or preoccupied with the thoughts of quitting its use, Continued excessive use of the Smartphone despite knowledge of having a persistent or recurrent psychological or physical problem as a result of its overuse); (2) Criterion B: the presence of ≥ 2 out of four functional impairment criteria assessed by replacing the words “Smartphone use” or SPU with “Instagram” (Excessive SPU resulting in recurrent psychological or physical problems, SPU in a physically dangerous, risky or hazardous situation (e.g., SPU while driving or crossing a road), or having other detrimental impacts on daily living, SPU causing impairment of school achievement, interpersonal relationships, or job performance, Excessive SPU leading to significant subjective distress, or is time-consuming and (3) exclusion criteria, whereby the BA is not better explained by an underlying psychiatric condition such as bipolar disorder or obsessive-compulsive disorder (Lin et al. 2016).

Additionally, by utilising fMRI, it is possible to illustrate *in vivo* brain activations in ‘Response to Reward’ conditions to better understand the neurobiology of this type of addiction (Sun et al. 2012; Kim et al. 2014; Zhang et al. 2016). Task-based fMRI employs Blood Oxygen Level Dependent Imaging (BOLD) to measure the hemodynamic response function (HRF) i.e., outshoot of oxygenated blood to local brain areas that are activated during tasks such as viewing pictures, videos, and decision-making in the cerebral cortex (Glover 2011; Sharifat et al. 2018; Lindquist et al. 2009; Huettel et al. 2014). This is particularly pertinent as according to the ‘Incentive sensitization theory of addiction’, attention bias and pathological motivation toward addiction-related cues such as pictures, movies, and words tended to elicit higher levels of arousal/ cravings in addicted individuals (Robinson and Berridge 2008).

Capra (1996), defined a paradigm as “a group of values, concepts, perceptions and practices that are shared by a community, which forms a unique vision of reality that is the foundation of the way a community organizes itself”. Paradigms are used to investigate individual differences, group behaviour, organizational behaviour, human factors, and cognitive behaviour among a group of subjects. In specific, an experimental fMRI paradigm refers to a temporal allocation of stimuli to obtain good BOLD activations by employing sensory cues e.g. emotional visual stimuli that are embedded in a block or event-related design to evoke HRF in the subjects (James et al. 2014). Designing a proper fMRI paradigm enables the researcher to achieve a good response accuracy and subsequently, identify regional brain activations in response to the stimulus being presented.

Increased craving or euphoria in an addicted subject can be studied by cue-reactivity paradigms using emotional visual cues to elicit feelings of arousal and craving during reward anticipation or ‘Response to Reward’ during reward delivery among the pathological users; as compared to the control group. It is one of the widely employed paradigms to evoke and evaluate craving for pathological gambling, Internet addiction, smartphone addiction, and pathological Instagram and Facebook usage (Chakraborty 2016; Sherman et al. 2018; Ko et al. 2009; Niu et al. 2016; Brand et al. 2016; Syed Nasser et al. 2019). A relatively new parameter with regards to the study of PIGU is the study of emotions. The exact nature of emotional motivational states in PIGU has not yet been adequately understood. Emotional stimuli are characterized by dimensions such as arousal, approach-avoidance (motivational direction) and emotional valence to describe the human perception of physical properties of the stimuli. Valence describes the feeling of pleasure in terms of positive or negative feelings evoked by the visual cues. Arousal is a physiological activity ranging from aroused/excited to unaroused/relaxed. Motivational direction refers to the directional aspect of the behaviour i.e., whether the respondent will decide to move towards/approach the stimuli or move away/avoid that stimuli (Feldman-Barrett et al. 2006; Mauss and Robinson 2009). Studies related to SNA have identified that the youth of today, namely young adults, tend to post ‘selfies’ on social media to gain peer approval and popularity via receiving ‘Likes’(Sherman et al. 2018). The uploaded risky and emotional negative posts-which often contain pictures taken from dangerous heights, rooftops, towers, near railway lines and on the road and photos with problematic interpersonal relationships such as with ex-boyfriends or ex-girlfriends-to receive ‘Likes’ from their peers (Kurniawan et al. 2017) and vice versa the evidence of risky lifestyle behaviours such as taking illicit drugs, alcoholism, eating disorders, etc. among the youth of today can be sought through data mining of publicly available Instagram posts (Zhou et al. 2017). Thus, we hypothesize risky and negative emotional pictures act as rewarding visual cues that can effectually modulate emotional experiences among PIGU in this age group. Hence, appropriate emotional cues for neuroimaging studies can be selected based on the ratings of these dimensional categories.

Validated flashcard sets or image databases are invaluable for a good cue-induced reactivity fMRI study. Although the availability of standardized databases such as the International Affective Picture System, and the Geneva Affective Picture Database that contain emotionally charged stimuli cannot be denied, there is no particular database on Smartphone-linked Instagram addiction; thus lacking appropriate images for potential future cue-induced fMRI studies to evaluate the ‘Response to Reward’ condition among smartphone addicts particularly in the domain of social networking applications such as Instagram.

To date, there is no local database with addiction-specific cues on PIGU that can elicit robust brain activations and portray similar neurobiological mechanisms as shown in SUDs. The main objective of this study is to validate a set of flashcards that can induce ‘Negative Emotional Valence’ and can potentially elicit brain activations in PIGU subjects. We aimed to create a database of ‘Neutral’ cues as a baseline and ‘‘Negative Emotional’ cues as the rewarding cues for future fMRI studies on PIGU. This validation study was designed and modified based on the method used by the Nencki Affective Picture System (NAPS)(Marchewka et al. 2014).

## Materials and Methods

We modified Sherman’s method on the assessment of the power of ‘Likes’ to evaluate the ‘Response to Reward’ condition employing negative emotional stimuli and ‘Neutral’ cues (Sherman et al. 2016). The pictures for this current study were selected from publicly available Instagram accounts after receiving approval from the account holders. The pictures were collected and classified into two categories (‘Negative Emotional’ cues and ‘Neutral’ cues). The former category included photographs such as ‘selfies’ taken from heights, taking photos while driving, loss of an interpersonal relationship and photos that were taken while doing dangerous and risky stunts. The latter category included pictures of landscapes, plants, trees, and object photographs in a greyscale background. Grayscale images provide very little information and elicit less interest compared to coloured images, hence it can reduce the involvement of the human visual system in extracting information from the images (Kather et al. 2017).

The initial classification of pictures into the selected categories was made by the principal team members (NSN, HS, AAR, SS). The selected images were then shown to 3 independent consultants (CSM, SIF, MFM), who gave their impartial opinion on the images by consensus. They acted as impartial judges to classify the pictures into 2 categories. Very bright, colourful pictures that provoked any positive emotions or a feeling of arousal were excluded under the ‘Neutral’ category. ‘Negative Emotional’ cues depicting photos taken in religious or sacred places were excluded to avoid any disputes. Any blurred photographs from both categories were excluded. At the beginning of the study, we had 300 pictures, out of which 100 ‘Neural” pictures and 100 ‘Negative Emotional’ pictures were selected. In approximately 99% of the cases, all of the judges classified the pictures into the same category (Cronbach’s α = .99). The finalized pictures (n=200), were then processed for the specifications of an fMRI paradigm. The pictures were resized and cropped to the specification of 600 ×400 with a resolution of 300 dpi using the freely available software, ‘Irfan View’ (sourced from https://www.irfanview.com/). The pictures were using JEPG file format.

## Participants

After acquiring ethical clearance from the local institutional committee (JKEUPM) (UPM/TNCPI/RMC/1.4.18.2), a validation study was conducted among 44 undergraduate students, 20 PIGU (16 girls, 4 boys; mean age 22.05 years) and 24 control subjects (18 girls, 6 boys; mean age 21.75 years) from Universiti Putra Malaysia. The subjects were recruited based on the evaluation of the online SAS-M and the modified IGAT questionnaires. Based on the recommended cut-off scores, subjects who attained a total score of ≥ 98 on SAS-M and ≥37 on the modified IGAT, along with the fulfillment of the diagnostic criteria proposed by Lin et al. (2016); were considered as PIGU. Conversely, those with a lower score from the cutoff values and who did not fulfill the diagnostic criteria were categorized as controls. Recruitment of participants for the study was made by advertising in the student portals and via communication with lecturers across all the faculties in Universiti Putra Malaysia (UPM). Responding participants were undergraduate students from various faculties in UPM and were selected by simple random sampling. Informed consent was obtained from all individual participants and the study abided by the rules advocated by the Declaration of Helsinki 1964.

## Rating scales and stimuli presentation

Before the commencement of the study, a briefing session was conducted among the study participants. The participants were instructed to answer the questions honestly and without discussing it with other people. The stimuli from both the categories were presented alternatively; with the limitation that no more than three stimuli from each category were presented in consecutive succession. The participants were instructed to view each picture for 3s and then respond to the 3 sub-questions related to emotional valence, motivation and arousal, for a duration of 3s each (see Appendix: Supplementary file 1).

We used an online picture evaluation method to rate our flashcards on a 9 point rating scale. The participant had to complete the sentence on the valence scale as “You are judging this image as …” (from 1 = very negative to 9 = very positive, with 5 = ‘Neutral’). Secondly, the participants judged the motivational direction by completing the sentence, “My reaction to this image is …” (from 1 = to avoid to 9 = to approach, with 5 = ‘Neutral’). Finally, participants judged the degree of arousal elicited by the pictures with the introductory sentence, “Confronted with this image, you are feeling: …” (From 1 = relaxed to 9 = aroused, with 5 = ‘Neutral’/ambivalent). The duration of the study was 45 minutes. The validation form was emailed to the participants and the responses were obtained via an online link. All responses were further analyzed using the statistical package SPSS version 22.

## Results

The ratings and descriptive statistics for the dimensions and pictures in each category by groups are presented in Table 1. PIGU and control groups showed differing emotional responses to the ‘Negative Emotional’ cues. Within the PIGU group, the ‘Negative Emotional’ cues elicited a positive feeling and they got aroused and preferred to approach those pictures. Whereas, the control group, although aroused by the ‘Negative Emotional’ cues, preferred to respond by avoiding those pictures. Both groups rated the ‘Neutral’ cues as pleasant and relaxing/un-arousing and were apt to approach those pictures.

**Table 1.** Descriptive statistics calculated separately for each dimension and category in the target groups

| Category |  | PIGU group |  |  | Control group |  |  |
| --- | --- | --- | --- | --- | --- | --- | --- |
|  |  | A | B | C | A | B | C |
| **‘Negative Emotional’** | **Mean±** | 5.005± | 4.83± | 5.60± | 4.445± | 4.304± | 5.697± |
|  | **SD** | 0.945 | 0.941 | 0.480 | 1.020 | 1.020 | 0.643 |
|  | **Min-Max** | 2.60-7.1 | 2.45-7.1 | 3.9-6.8 | 2.67-7.21 | 2.5-7.08 | 3.83-7.13 |
|  | **N** | 100 | 100 | 100 | 100 | 100 | 100 |
| **‘Neutral’** | **Mean±** | 6.165± | 6.011± | 4.765± | 6.292± | 6.294± | 4.043± |
|  | **SD** | 0.613 | 0.607 | 0.452 | 0.639 | 0.684 | 0.636 |
|  | **Min-Max** | 3.7-7.5 | 3.5-7.35 | 3.65-5.7 | 4.08-7.63 | 3.96-7.58 | 2.83-5.71 |
|  | **N** | 100 | 100 | 100 | 100 | 100 | 100 |
| **‘Negative Emotional’ and ‘Neutral’ combined** | **Mean±** | 5.585± | 5.420± | 5.187± | 5.369± | 5.299± | 4.87± |
|  | **SD** | 0.986 | 0.987 | 0.629 | 1.256 | 1.322 | 1.046 |
|  | **Min-Max** | 2.60-7.5 | 2.45-7.35 | 3.65-6.8 | 2.67-7.63 | 2.5-7.58 | 2.83-7.13 |
|  | **N** | 200 | 200 | 200 | 200 | 200 | 200 |

SD: Standard Deviation; A: Valence; B: Approach-Avoidance; C: Arousal; Min-Max: Minimum-Maximum value; N= Number of pictures

## Correlation analyses of emotional dimensions for the groups and categories

Taking the pictures as cases, we conducted a correlation analyzing using Pearson’s correlations to examine the relationships between ratings of valence, arousal, and approach-avoidance for each category and group separately; as described in Table 2 and shown in Figure 1, 2 and 3 respectively. All dimensions were highly correlated in both the PIGU group and the control group (p <0.001) as demonstrated in Table 2. The control group, however, had a strong negative correlation compared to the PIGU group in all cases, except for the correlations between approach-avoidance and arousal, in which both groups exhibited a strong positive correlation. Correlation coefficients of the control group were also directly compared to those of the addicted group using a calculation for the test of the difference between two independent correlation coefficients (Preacher, 2002). This calculation involves converting the two correlation coefficients into z scores using Fisher’s r-to-z transformation. Then, making use of the sample size employed to obtain each coefficient, these z scores are compared using Formula 2.8.5 from Cohen and Cohen (1983, p.54):

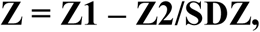

**Table 2.** Pearson’s correlation of ratings of valence, arousal and approach-avoidance for each category in the target groups

| Category | Dimension | PIGU group<br>Pearson’s ‘r’ |  | Control group<br>Pearson’s ‘r’ |  |
| --- | --- | --- | --- | --- | --- |
|  |  | C | A | C | A |
| ‘Negative Emotional’ | A | -.372* |  | -.795* |  |
|  | B | -.423* | .971* | -.806* | .985* |
| ‘Neutral’ | A | -.716* |  | -.895* |  |
|  | B | -.745* | .944* | -.905* | .980* |

A: Valence; B: Approach-Avoidance; C: Arousal
\*Correlation is significant at $p < 0.05$

**Figure 1.**
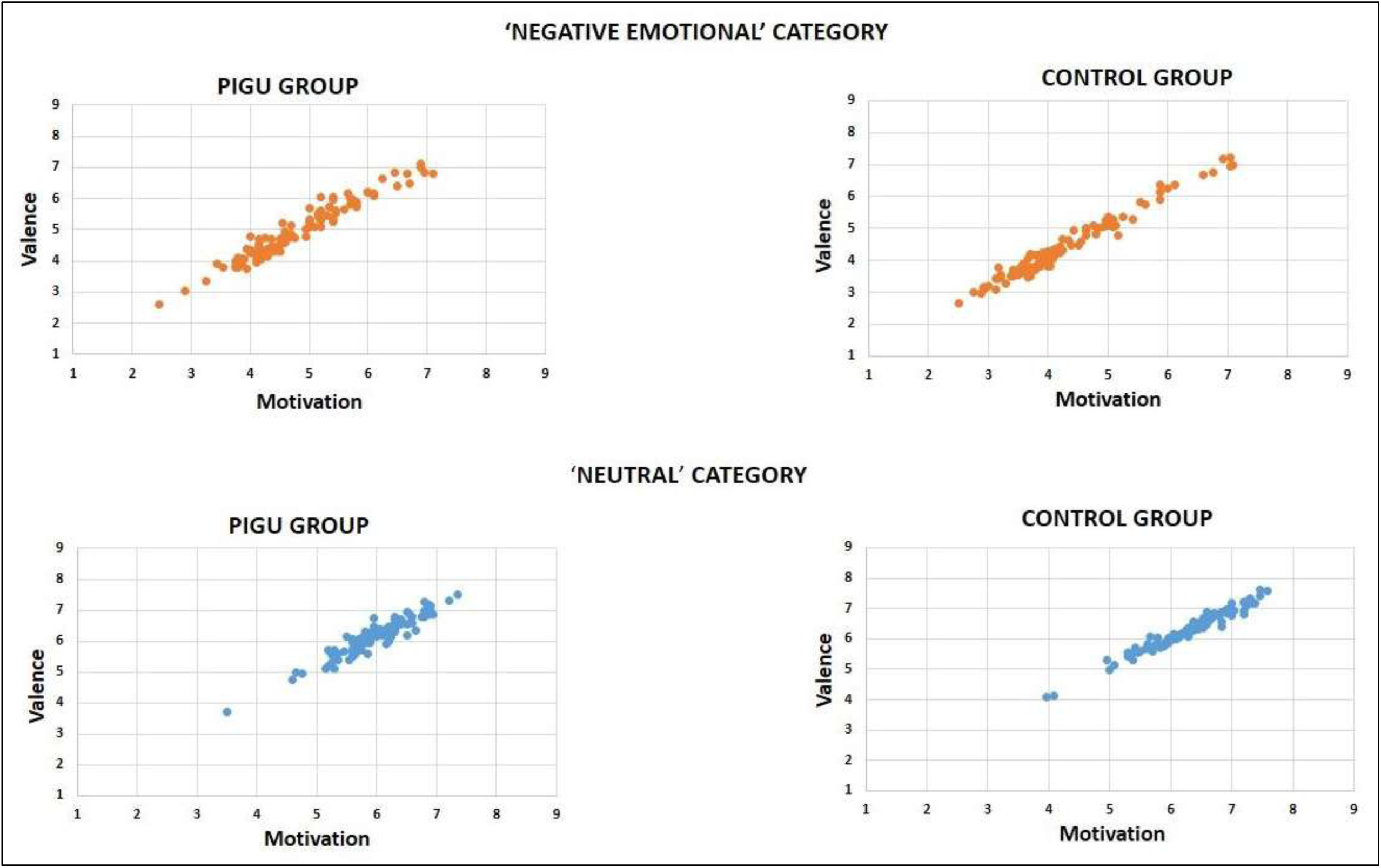
Scatter plot chart depicting the relationship between scores by PIGU and control groups for the two different image categories showing behavioral ratings for emotional valence (y-axis) and motivation (x-axis) in each category for the PIGU and control groups. *Every single dot represents the mean rating for a particular picture on a two-dimensional scale.

**Figure 2.**
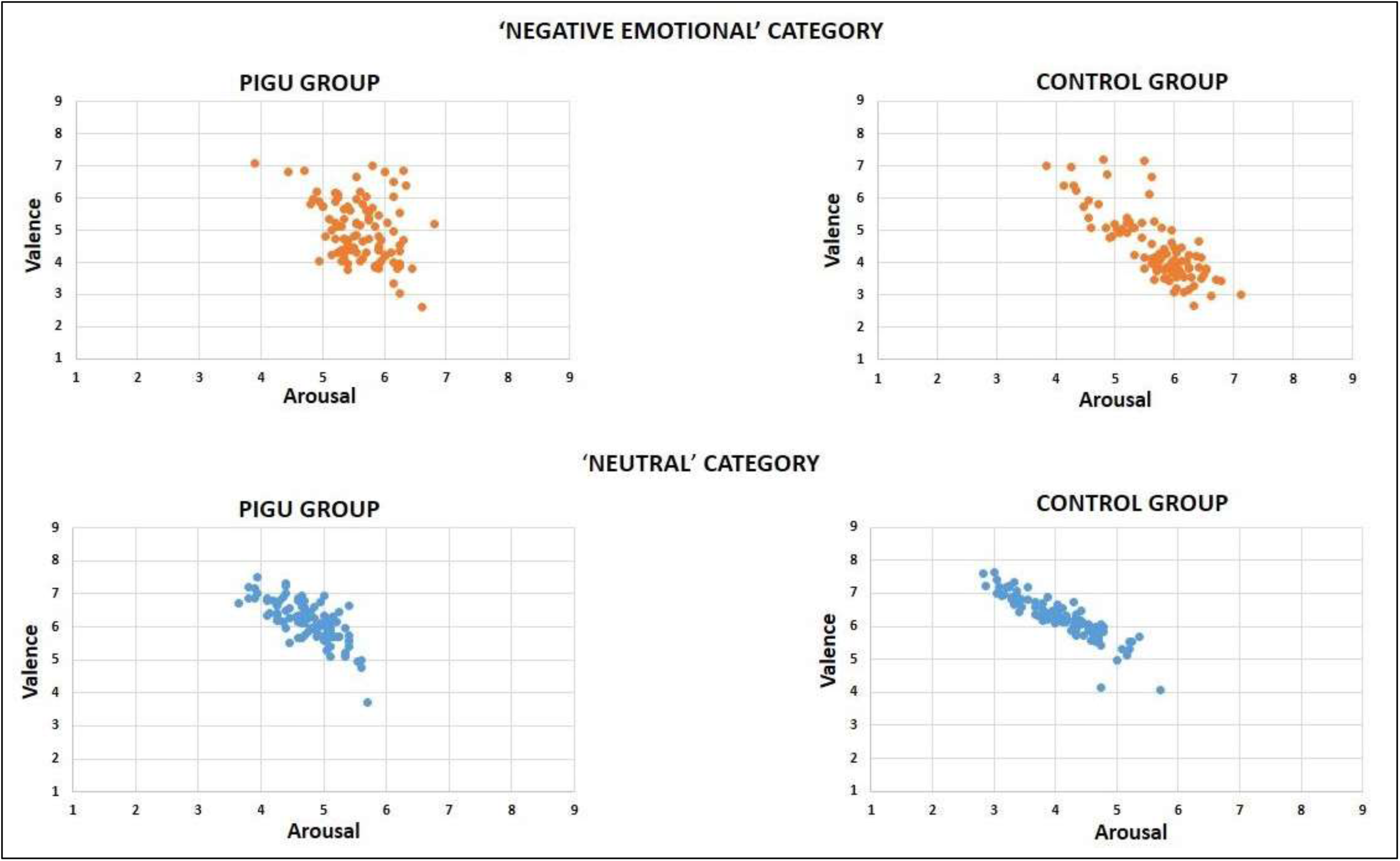
Scatter plot chart depicting the relationship between scores by PIGU and control groups for the two different image categories showing behavioral ratings for emotional valence (y-axis) and arousal (x-axis) in each category for the PIGU and control groups. *Every single dot represents the mean rating for a particular picture on a two-dimensional scale.

**Figure 3.**
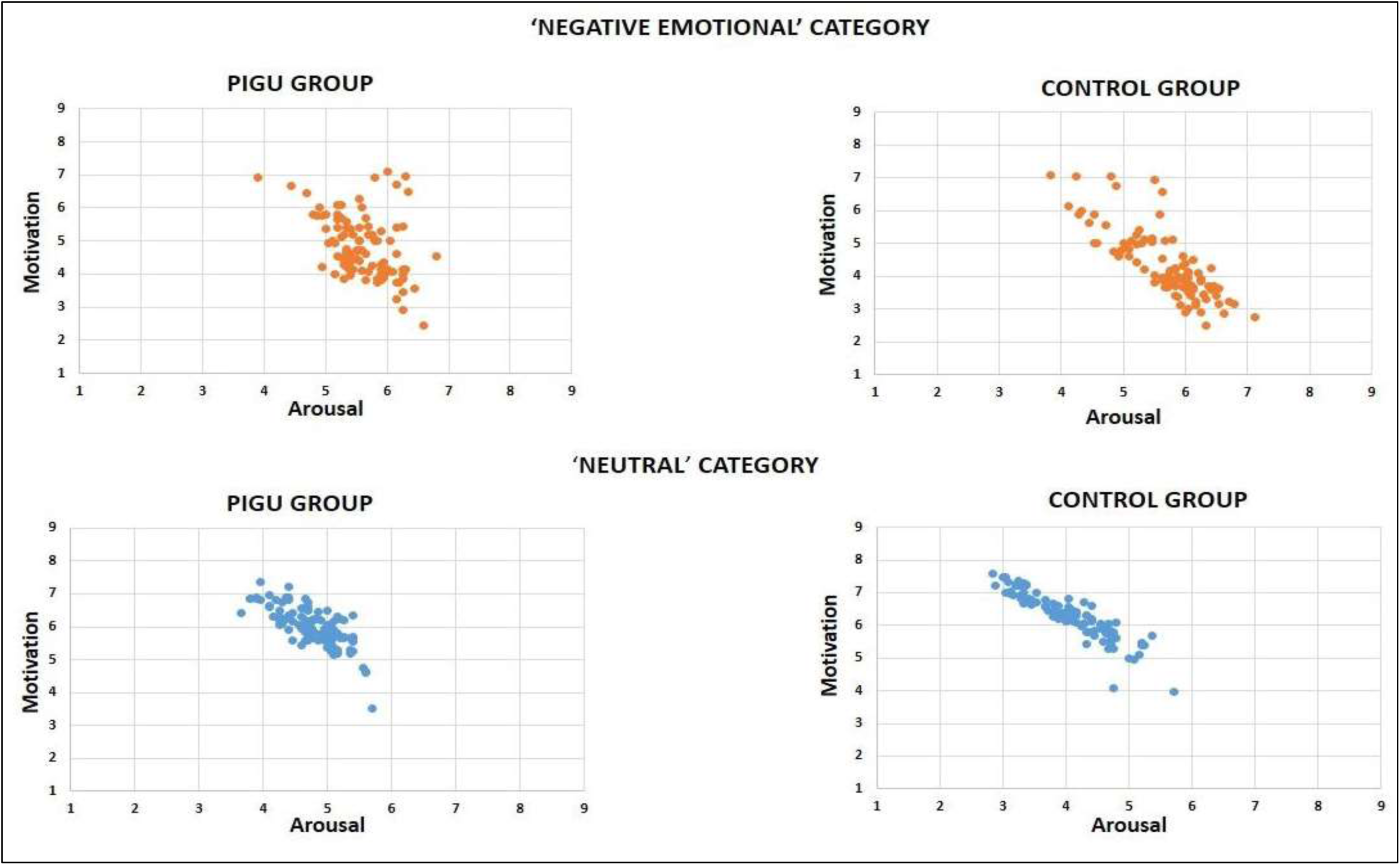
Scatter plot chart depicting the relationship between scores by PIGU and control groups for the two different image categories showing behavioral ratings for motivation (y-axis) and arousal (x-axis) in each category for the PIGU and control groups. *Every single dot represents the mean rating for a particular picture on a two-dimensional scale.

Where, 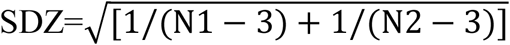, and N1 and N2 are the sample sizes. In case of the correlation between emotional valence and arousal, the differences between the PIGU group and control group for the ‘Negative Emotional’ category showed a strong positive effect (z = 4.834, p < .001, effect size = 0.69) compared to (control group-PIGU group), which showed a strong negative effect (z=−4.83). Similarly, in the case of arousal correlated with approach-avoidance, the differences between the PIGU group and control group (PIGU group-control group) for the ‘Negative Emotional’ category, showed a strong positive correlation (z = 4.625, p <0.001, effect size = 0.66) compared to (control group-PIGU group), which showed a strong negative correlation (z=−4.625). For the condition of emotional valence correlated with approach-avoidance differences between the (control group-PIGU group) for the ‘Negative Emotional’ category showed a medium positive correlation (z = 2.32, p <0.001, effect size = 0.33) compared to (PIGU group-control group), which showed a medium negative correlation (z=−2.32).

## Discussion

This study focused on developing and validating visual cues called the ‘Problematic Instagram Use Flashcards’, with the focus on ‘Negative Emotional Valence’ cues to evoke an emotional response to risk-taking behaviour among the problematic Instagram users in the young adults’ age group. Conversely, the ‘Neutral’ cues acted as a baseline stimulus to be used as addiction-specific cues in potential future fMRI studies.

In the process of creating this database, we focused on dividing the cues into 2 categories -‘Negative Emotional’ and ‘Neutral’ cues. The ‘Negative Emotional’ cues included images denoting problematic use of Instagram; such as pictures taken from high altitude, texting while driving, dangerous stunts and loss of an interpersonal relationship, which the PIGUs could perhaps relate to and deem as arousing to them. ‘Neutral’ cues involving nature and objects in grayscale and dull-coloured background were used as baseline stimuli. All the pictures in the present database were of good quality with a resolution of 300 dpi. We determined the correlations between emotional valence, arousal, and approach-avoidance for the ‘Negative Emotional’ and ‘Neutral’ categories, as scored by the PIGU and control groups.

The results suggest that there is a distinct profile for PIGU group with regards to their experience of emotions when presented with ‘Negative Emotional’ cues that we deem as addiction-specific cues. Taking into account the differences in the correlation coefficients between the 2 groups, together with the scatter plot charts, correlations between arousal and emotional valence, arousal and avoidance-approach were strongly positive in the PIGU group as compared to the control group for the ‘Negative Emotional’ category cues. We found that PIGU subjects compared to the controls were vulnerable to addiction-specific cues and were more frequently aroused by ‘Negative Emotional’ type of stimuli as they frequently used such type of images to receive more ‘Likes’ on Instagram. When comparing between emotional valence and motivation among the controls, the pictures in the ‘Negative Emotional’ category were rated as unpleasant. Hence, they had perceived it as ‘negative’ and would prefer to ‘avoid’ these cues. In contrast to this, the PIGU subjects had abnormal perception and rated these stimuli to be pleasant. We postulate this to be a result of the abnormally accentuated need to gain popularity and acceptance among their peers that has forced the PIGU subjects to strongly indulge in risk-taking behaviours and had increased their motivation to seek risky situations. These findings are in line with the study by Volkow et al. (2010) that observed a greater activation of the motivation circuit upon exposure to drug cues, ultimately resulting in the compulsive intake of illicit drugs. In the latter study, addicted participants showed a tendency towards approaching the ‘Negative Emotional’ cues.

Next, when studying arousal compared with emotional valence, we found that ‘Neutral’ category cues did not elicit any excitement or arousal in both the PIGU and the control groups. Interestingly, when presented with ‘Negative Emotional’ cues, both groups reported increased ratings for arousal. This finding is consistent with the studies on Internet addiction and alcohol addiction, which reported non-addicts can also get aroused in response to the addictive cues (Sinha and Li 2007; Niu et al. 2016). This may be due to the daily use of the social networking application or closely associating this kind of behaviour with themselves, friends or family members (Niu et al. 2016).

Although both groups were aroused during the ‘Negative Emotional’ cues, we observed that the mean rating of arousal in the control group was greater than that of the PIGU subjects. This is because the controls are not frequently exposed to risk-taking behaviours. Thus, when confronted with ‘Negative Emotional’ cues, which are considered “taboo”, socially unaccepted and should be discouraged, they found it exciting (Duncan and Petosa 1994). Nevertheless, they correctly responded by avoiding such cues. As anticipated, the PIGU subjects were also aroused by these images and tended to respond by approaching these cues. Nevertheless, a few participants showed reduced arousal, which we speculate may be due to their over-involvement in extreme levels of risk-taking behaviour leading to the development of tolerance. Because of their chronic exposure to such content in their Instagram account, the PIGU group are postulated to become very aroused to extremely ‘Negative Emotional’ kind of images and will approach the ‘Negative Emotional’ cues.

Lastly, the age difference has a significant role in the processing of emotional stimuli. Age can also influence the degree of arousal and hence can affect brain activation in fMRI studies. Previous studies have found that older people, middle-aged people and younger adults differ in perception and processing of emotional stimuli. A study by Grühn and Scheibe (2008) found older people compared to the young adults over-rate emotional valence and arousal dimensions (Grühn and Scheibe 2008). Whereas, Gilet et al (2012) reported middle-aged people showed intense mean ratings for positive pictures compared to older people (Gilet et al. 2012). Thus, to avoid any bias and disputes, we limited the study to undergraduate students of a similar age group in a public university in Malaysia.

These results suggest that PIGU can produce long term alterations in the way the users experience emotions when presented with ‘Negative Emotional’ category cues. The decision-making towards ‘Negative Emotional’ cues was “stronger” among the control group to guide them towards adaptive objectives, rather than towards ‘Negative Emotional’ cues as observed among the PIGU subjects. Furthermore, neuroimaging studies have highlighted the role of emotional markers in decision-making (Bechara 2004; Bechara et al. 2002). The prefrontal cortex (PFC) is a key brain area responsible for decision-making to find solutions in a given task and decides whether to ‘go’ towards or to ‘inhibit’ the stimulus. In agreement with these studies, the decision-making in the PIGU group was affected when presented with ‘Negative Emotional’ cues due to inability to exhibit an inhibitory response to such stimuli (Syed Nasser et al. 2020). This study helps to explain why the ‘Negative Emotional’ cues associated with Instagram usage increases the motivational behaviour of the PIGU group towards such stimuli as opposed to the control group. This inability to regulate one’s control of negative behaviour may lead to a distorted perception of social norms.

From a set of 100 pictures in each category, we finally selected 20 ‘Negative Emotional’ and 20 ‘Neutral’ pictures. The suitable ‘Negative Emotional’ pictures were selected on the basis that the pictures elicited a positive feeling and arousal and a tendency to approach among the PIGU subjects, while the same pictures rated as negative and induced a tendency to avoid among the control group. The pleasant, un-arousing pictures in both groups were selected as suitable ‘Neutral’ stimuli. The appropriate cues selected based on this validation study will enable the researchers to identify the regional brain activation in the mesocorticolimbic system in future fMRI studies.

This study focused specifically on the risk-taking behaviours of undergraduate students who are in the young adults’ age group, hence it is likely that the behavioural influence may vary among adolescents as reported by Sherman et al. (2018) and older adults, who may be preoccupied with different types of social norms and lifestyle behaviours. We recommend that future studies should design their paradigms to facilitate adolescents and adults who may require a different set of flashcards for an fMRI study that is targeted to age-specific content. Another limitation of this study is the small sample size and limited theme of flashcards stimuli. Further validation of this picture database employing computerized SAM or the slider scale methodology is recommended. Additionally, as we had conducted an online survey, we are unsure regarding the exact duration of time the subjects took to answer each question. An additional minor limitation of this present study is the lack of very negative (low emotional valence) pictures with a high avoidance-approach outcome.

## Conclusion

A validated database of flashcard images on problematic Instagram usage has been created to potentiate future fMRI ‘Response to Reward’ cue-induced reactivity studies among young Malaysian adults. The database offers high-resolution addiction-specific cues that could potentially activate the mesocorticolimbic reward system of the brain when performing fMRI. Images from our database will be made available to interested researchers upon request made to the corresponding author.

## Data Availability

The data that support the findings of this study are available upon reasonable request from the corresponding author.

## Acknowledgements

This study was funded by Universiti Putra Malaysia research grants, Geran Putra (GP-IPS/2017/9580800 and GP/2017/9549800) awarded to Associated Professor Dr. SubapriyaSuppiah. All the authors declare that there is no conflict of interest in the publication of this research. The authors would like to thank the students from different faculties of UPM who participated in this study.

## Appendix

**Supplementary file 1.**
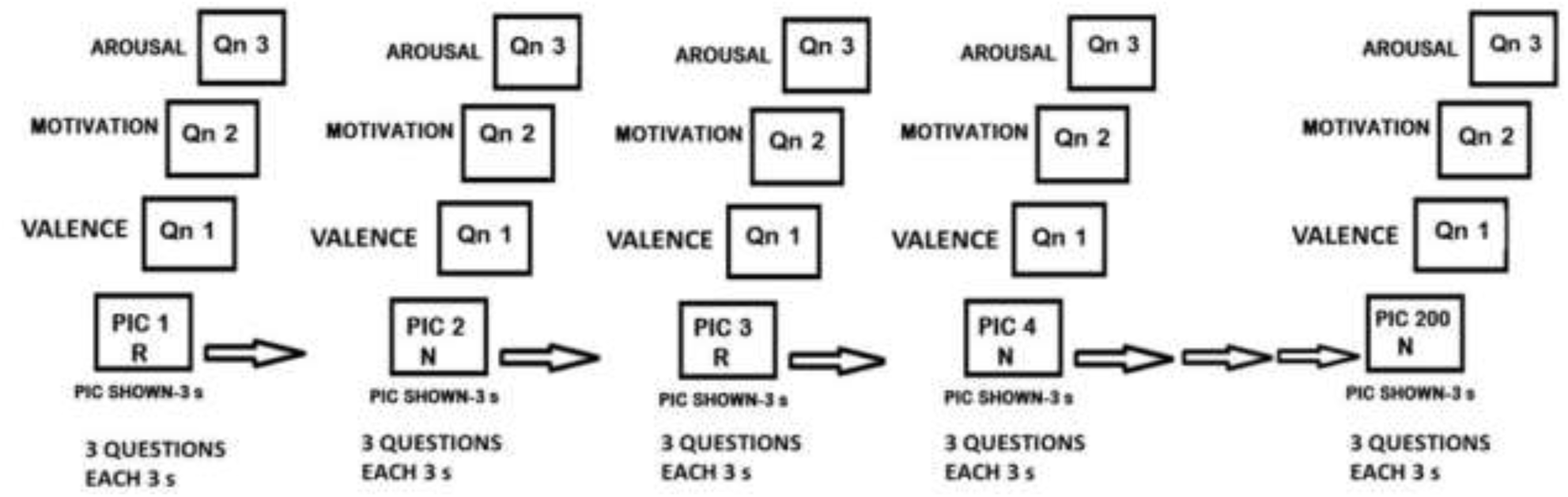
Stimuli presentation -the off-scanner paradigm

